# Clinical predictors of treatment outcome in Melanesian adults with Tuberculous Meningitis at the Kundiawa General Hospital in Papua New Guinea

**DOI:** 10.1101/2022.09.03.22279565

**Authors:** Stanley Aiwara, Izzard Aglua

## Abstract

**Background:** Tuberculous meningitis is the most severe form of extrapulmonary TB and accounted for 5% of 10 million global TB cases in the WHO 2018 report, with mortality as high as 19% in children and 30% in adults. Clinicians in resource-poor settings are often challenged by limited diagnostic and therapeutic options for optimal patient care, and often rely on clinical parameters for diagnosis, treatment, monitoring and outcome.

**Aim:** This study was done to identify potential clinical predictors of i) 28-day mortality and ii) length of hospitalization, amongst Melanesian adults with TB meningitis at a provincial hospital in Papua New Guinea.

**Method:** A retrospective observational study was conducted on 65 Melanesian adults with TB meningitis at a provincial hospital in Papua New Guinea between 2015 and 2019.

**Result:** High case fatality (49%) and mortality rates (2.22 per 100 000 per year) for TB Meningitis were observed in this study. Even higher case fatality of 93% observed for HIV-TBM co-infection. 28-day mortality associated with HIV-TBM co-infection (p-value=0.007, 95% CI 2.49-289.19), positive fluid balance 24-hours after admission (p-value=0.019, 95% CI 1.23-10.19) and admission GCS ≤10 (p-value=0.049, 95% CI 1.01-16.58).

**Conclusion:** Study showed high case fatality (49%) and mortality rates (2.22 per 100,000 per year) for TB Meningitis in Melanesian adults, with significantly high case fatality (93%) for HIV-TBM co-infection. HIV-TBM co-infection was strongly associated with 28-day mortality whilst a positive fluid balance 24-hours after admission and admission-GCS ≤10 were weakly associated with 28-day mortality.

## Background

Tuberculous Meningitis (TBM) is the most severe form of extrapulmonary Tuberculosis (TB) which accounted for 5% of 10 million global TB cases reported by WHO in 2018, with a high mortality and morbidity ^1, 2^. TB meningitis studies have reported mortalities as a high as 19% in children and 30% in adults, with an associated high disease burden of neurological disability after treatment ^3-5^. Mortality was as high as 80% in studies from Sub-Saharan Africa and Asia ^5^.

The ongoing challenge for effective treatment with optimal outcome is further compounded by limited diagnostic and therapeutic options for timely and appropriate treatment in resource-constrained settings ^6-7^. Clinicians in these settings often rely on clinical parameters for diagnosis, treatment, monitoring and outcome prediction.

This observational study was done to explore the potential role of certain clinical and cerebrospinal fluid (CSF) parameters in predicting treatment outcome amongst Melanesian adults with TB Meningitis at a Provincial Hospital in Papua New Guinea. The primary outcome measure was death or discharge at 28 days, with length-of-hospitalization as a secondary outcome. Potential predictors of outcome assessed included duration of illness, admission Glasgow Coma Score (GCS), mean arterial pressure (MAP) on admission, plasma sodium on admission, fluid-balance status 24-hours after admission, HIV-TBM co-infection and CSF findings (polymorphs, lymphocytes and acid-fast bacilli), besides age and gender^8, 9^.

## Aim

To explore for potential clinical predictors of i) death at 28 days and ii) length of in-hospital stay, amongst Melanesian adults with TB meningitis in a resource-limited setting in Papua New Guinea.

## Method

A retrospective observational study was conducted on a total of 65 Melanesian adults with TB meningitis at the medical ward of the Kundiawa General Hospital in Papua New Guinea between 2015 and 2019.

A modified Thwaites Criteria using clinical and investigative findings was employed for the diagnosis of Tuberculous Meningitis ^10, 11^, whereby a patient with at least four of the following thematic features was diagnosed as a case of TBM:

Meningitis Features

i. Symptom (headache, fever, photophobia, vomiting, fitting) ≥ 2 weeks
ii. Sign (Neck stiffness ± Kernig’s or Brudzinski’s sign)
iii. Altered conscious status
iv. Focal neurological symptom
v. Positive CSF finding (positive Acid-Fast Bacilli, lymphocyte predominance, elevated glucose with low protein).

Tuberculosis Features

i. History (positive contact, family history)
ii. Symptoms (chronic productive cough, night sweats, weight loss, hemoptysis)
iii. Sign (wasting, regional lymphadenopathy, extrapulmonary TB sign)
iv. Test (consistent chest radiograph, sputum positive, positive CSF finding)

Of the four thematic features required, at least one of the five Meningitis features and one of the four Tuberculosis features was required to constitute a clinical or bacteriological diagnosis of Tuberculous Meningitis. Focal neurological symptoms included cranial nerve palsies. CSF acid-fast bacilli (AFB) was assessed by polymerase chain reaction (PCR).

Study data was collected from patients’ information chart using a semi-structured document review form by two investigators at a monthly interval over a 5-year period. Raw data was entered onto an excel spread sheet and imported into Stata 15.1 for analysis. Summary statistics and regression analysis (univariate & multivariate) were performed on variables of interest for possible relationships between predictors and outcome. Variable selection for analysis was guided by relevant literature indication and clinical observation of potential influence on meningitis (chronic/acute) outcome. Logistic regression was applied for 28-day mortality given its dichotomous nature, whilst liner regression was applied for length-of-hospitalization for its numeral nature. The primary outcome measure was death at 28 days, and secondary measure was length-of-hospitalization. Parameters assessed included duration of illness on admission (days), admission Glasgow Coma Score (GCS), mean arterial pressure (MAP) on admission, plasma sodium (Na^+^) on admission, fluid balance status 24-hours after admission, HIV-TBM co-infection and CSF findings (polymorphs, lymphocytes and AFB), besides age and gender ^8,9^. The retrospective nature of the study limited its ability to effectively asses neurological deficit (neuromotor, cognitive) as an outcome measure, owing to data insufficiency.

65 patients, over 13 years of age and diagnosed with TB meningitis (per criteria) with or without HIV infection were included for this study. All study subjects received a course of dexamethasone (2-8 mg iv/oral/NGT) during in-hospital treatment.

## Result

**▪ 28-Day Mortality**

^*\**^*Following results pertain to table 1*.

**Table 1.** showing summary data with result of regression analyses for 28-day mortality.

|  |  |  |  |  | Univariate Analysis |  |  | Multivariate Analysis |  |  |
| --- | --- | --- | --- | --- | --- | --- | --- | --- | --- | --- |
| Variable |  | Discharged | Died | N<br>(65) | OR | 95% CI | p-value | aOR | 95% CI | p-value |
| Gender | Female | 15 | 12 | 27 |  |  |  |  |  |  |
|  | Male | 18 | 20 | 38 | 1.39 | 0.52-3.74 | 0.516 | 2.36 | 0.46-11.98 | 0.301 |
| Age | <18 | 2 | 1 | 3 | (ref) |  |  | (ref) |  |  |
|  | 18-37 | 25 | 20 | 45 | 1.6 | 0.14-18.94 | 0.709 | 0.44 | 0.019-9.84 | 0.602 |
|  | 38-57 | 4 | 7 | 11 | 3.5 | 0.24-51.90 | 0.363 | 1.25 | 0.054-28.80 | 0.889 |
|  | >57 | 2 | 4 | 6 | 4 | 0.21-75.66 | 0.355 | 1.38 | 0.037-51.21 | 0.86 |
| Admission GCS | 15 | 11 | 5 | 16 | (ref) |  |  | (ref) |  |  |
|  | 11-14 | 15 | 14 | 29 | 2.05 | 0.57-7.41 | 0.272 | 2.42 | 0.36-16.30 | 0.365 |
|  | ≤10 | 7 | 13 | 20 | 4.09* | 1.01-16.58 | 0.049 | 3.04 | 0.36-25.51 | 0.306 |
| HIV Status | Negative | 32 | 20 | 52 |  |  |  |  |  |  |
|  | Positive | 1 | 12 | 13 | 19.2* | 2.32-159.18 | 0.006 | 26.81* | 2.49-289.19 | 0.007 |
| Illness Duration (Weeks) | 2 – 4 | 20 | 18 | 38 | (ref) |  |  |  |  |  |
|  | > 4 - 12 | 5 | 9 | 14 | 1.5 | 1.02-15.3 | 0.257 |  |  |  |
|  | > 12 | 8 | 5 | 13 | 3.01 | 0.54-19.03 | 0.36 |  |  |  |
| Admission Systolic BP (mmHg) | 100-140 | 28 | 27 | 55 | (ref) |  |  |  |  |  |
|  | <100 | 1 | 0 | 1 | - |  |  |  |  |  |
|  | >140 | 4 | 5 | 9 | 1.3 | 0.31-5.35 | 0.72 |  |  |  |
| Fluid Balance | Negative | 25 | 15 | 40 |  |  |  |  |  |  |
|  | Positive | 8 | 17 | 25 | 3.54* | 1.23-10.19 | 0.019 | 4.11* | 0.93-18.13 | 0.062 |
| Admission Na<sup>+</sup> | Normal | 7 | 7 | 14 |  |  |  |  |  |  |
|  | Abnormal | 26 | 25 | 51 | 0.962 | 0.29-3.14 | 0.948 |  |  |  |
| CSF Polymorphs | Absent | 8 | 2 | 10 | (ref) |  |  | (ref) |  |  |
|  | Present | 18 | 18 | 36 | 4 | 0.74-21.50 | 0.106 | 3.05 | 0.29-32.16 | 0.353 |
|  | No Result | 7 | 12 | 19 | 6.86* | 1.12-41.83 | 0.037 | 1.17 | 0.021-62.94 | 0.939 |
| CSF Lymphocytes | <10 | 18 | 13 | 31 | (ref) |  |  |  |  |  |
|  | >10 | 8 | 7 | 15 | 1.21 | 0.35-4.19 | 0.762 |  |  |  |
|  | No Result | 7 | 12 | 19 | 2.37 | 0.73-7.68 | 0.149 |  |  |  |
| Gram Stain (CSF) | Negative | 23 | 15 | 38 | (ref) |  |  | (ref) |  |  |
|  | Positive | 2 | 2 | 4 | 1.53 | 0.19-12.09 | 0.685 | 0.37 | 0.033-4.21 | 0.425 |
|  | No Result | 8 | 15 | 23 | 2.88 | 0.98-8.44 | 0.055 | 3.08 | 0.11-84.54 | 0.506 |
| Gene X-pert (CSF) | Negative | 20 | 16 | 36 | (ref) |  |  |  |  |  |
|  | Positive | 5 | 3 | 8 | 0.75 | 0.16-3.62 | 0.72 |  |  |  |
|  | No Result | 8 | 13 | 21 | 2.03 | 0.68-6.10 | 0.206 |  |  |  |
| **TOTAL** |  | **33** | **32** | **65** |  |  |  |  |  |  |

### *Case fatality rate* for TB meningitis was 49% (32/65)

Mortality rate was 2.22 per 100 000 adults per year in the province as at 2019 population data (adult population 278, 971) ^12^. For an estimated population of 400 000 adults, 9 would have died from TB meningitis every year, at the given mortality rate (2.22 per 100 000 per year) for the 5-year study period.

### HIV-TBM coinfection

A statistically significant, p-value=0.006, 95% CI 2.32-159.18, association between HIV-TBM coinfection and 28-day mortality observed on the univariate regression, with an odds ratio of 19.2. This association remained significant, p-value=0.007, 95% CI 2.49-289.19, after controlling for confounders in multivariate regression. 93% (12/13) of HIV-positive TBM patients died at 28 days whilst 38% (20/52) of HIV-negative TBM patients died at 28 days in hospital. Hence, case fatality rate of 93% for HIV-positive patients with TBM.

### Fluid Balance

A significant association was observed between a positive fluid-balance 24-hours after admission, p-value= 0.019, 95% CI 1.23-10.19, and 28-day mortality (odds ratio 3.54) which was marginally significant, p-value=0.062, 95% CI 0.93-18.13, in the multivariate model. 68% (17/25) of TBM patients with a positive fluid balance in the first 24-hours of admission died, while 37.5% (15/40) of TBM patients with a negative fluid balance died at 28 days.

### Admission GCS

A significant association was also observed between admission GCS ≤ 10 and 28-day mortality, p-value=0.049, 95% CI 1.01-16.58, OR 4.09, but was not significant in multivariate model, p-value=0.306, 95% CI 0.36-25.51. 65% (13/20) of TBM patients with GCS ≤ 10 on admission died at 28 days, giving a case fatality rate of 65%. 30% (20/65) of study subjects had GCS ≤ 10 on admission.

^*^There was no significant association observed between 28-day mortality and the other variables analyzed.

**▪ Length of Hospitalization**

## Discussion

The case fatality (49%) and mortality (2.22 per 100 000 per year) rates for adult Tuberculous Meningitis observed in this study are comparably higher than that seen in similar studies from resource-limited settings around the world ^13-15^. Apart from diagnostic and therapeutic limitations, biological determinants inherent to *mycobacterium tuberculosis* like virulence and drug-resistance, together with the site and nature of infection, may account for similar higher mortalities for TB meningitis also observed in some resource-rich settings ^16-18^.

### HIV-TBM Co-infection

HIV-TBM Co-infection was observed to be an independent predictor for 28-day mortality in this study, agreeing with similar studies comparing mortality between HIV-positive and HIV-negative patients with TB meningitis ^19-21^. Although a low incidence of 16.92% in this study, HIV-TBM co-infection was associated with a significantly high case fatality of 93% and a statistically significant association with 28-day mortality even after controlling for confounding in the multivariate model (p-value 0.007, 95% CI 2.49-289.19).

### Fluid Balance

A positive fluid balance 24-hours after admission was also associated with 28-day mortality, p-value=0.019, 95% CI 1.23-10.19. However, the association was marginally significant in the multivariate regression (p-value=0.062, 95% CI 0.93-18.13). This may be owing to either confounding or a type two error (β) from a small sample size of 65. This study may have been underpowered which statistically reduces the strength and measure of the association between the variable (positive 24-hour fluid balance) and 28-day mortality. Hence, larger prospective cohorts would be ideal to further explore and/or clearly ascertain the association between a positive fluid balance and mortality in TBM patients.

Nevertheless, this result highlights the need for optimal management of fluid and electrolyte balance during in-patient treatment of TB meningitis with associated raised intracranial pressure to improve mortality ^22, 23^. A positive fluid balance 24-hours after admission should also alert clinicians to possible onset or progression of acute renal insufficiency to failure which will inadvertently contribute to increased mortality. Raised intracranial pressure from an infectious brain insult like TB Meningitis may also induce syndromes of salt wasting or excess anti-diuretic hormone action (SIADH) with resulting hyponatremia and fluid depletion or repletion respectively, which may in turn entail functional neurological derangement with poor outcomes ^24-27^.

### Admission GCS

65% (13/20) of TBM patients with admission GCS ≤ 10 died at 28 days. A significant association between admission GCS ≤ 10 and 28-day mortality was seen in the single-variable regression (p-value=0.049, 95% 95% CI 1.01-16), but not in the multivariate model (p-value=0.306, 95% CI 0.36-25.51). This may also be owing to an underpowered study of small sample size (n=65) or confounding. Nevertheless, depressed conscious state has long been identified as an independent prognostic factor for poor outcomes in Tuberculous, and other forms of chronic meningitis around the world ^28^. Delayed presentation to health care facilities with late treatment initiation in developing countries continue to influence disease severity and conscious level on admission, as reflected in this study by a high proportion (30%) of patients with lower admission GCS, and subsequent poor outcomes ^29,30^. Whilst GCS does not qualify as an independent predictor for 28-day mortality from this analysis, it is undoubtedly a significant clinical indicator of progress and outcome in TBM patients.

### Duration of Hospitalization

The duration of hospitalization ranged from 1 to 473 days, with a mean duration of 36.62 days. 90% of subjects stayed more than two weeks in the hospital (*table 2, figure 1*), and 46% (*figure 1*) stayed more than a month. These highlight the ongoing challenge for resource considerations with longer in-hospital care for chronic meningitis patients in resource-poor settings.

**Table 2.** showing summary statistics for duration of hospitalization.

| Length of Hospitalization |  |  |
| --- | --- | --- |
|  | Percentile<br>s | Day<br>s |
|  | 1% | 1 |
|  | 5% | 1 |
|  | 10% | 2 |
|  | 25% | 6 |
|  | 50% | 26 |
|  | 75% | 43 |
|  | 90% | 59 |
|  | 95% | 119 |
|  | 99% | 473 |
| Observations | 65 |  |
| Mean | 36.62 |  |
| Std. Deviation | 63.04 |  |

- Range: 1 day to 473 days. - Mean duration of admission: 36.62 days, SD 63.04 - Cumulative Frequency: 50% < 26 days, 90% < 59 days.
\*There was no statistically significant association observed between any of the assessed variables and length of hospitalization in the regression models.

**Figure 1.**
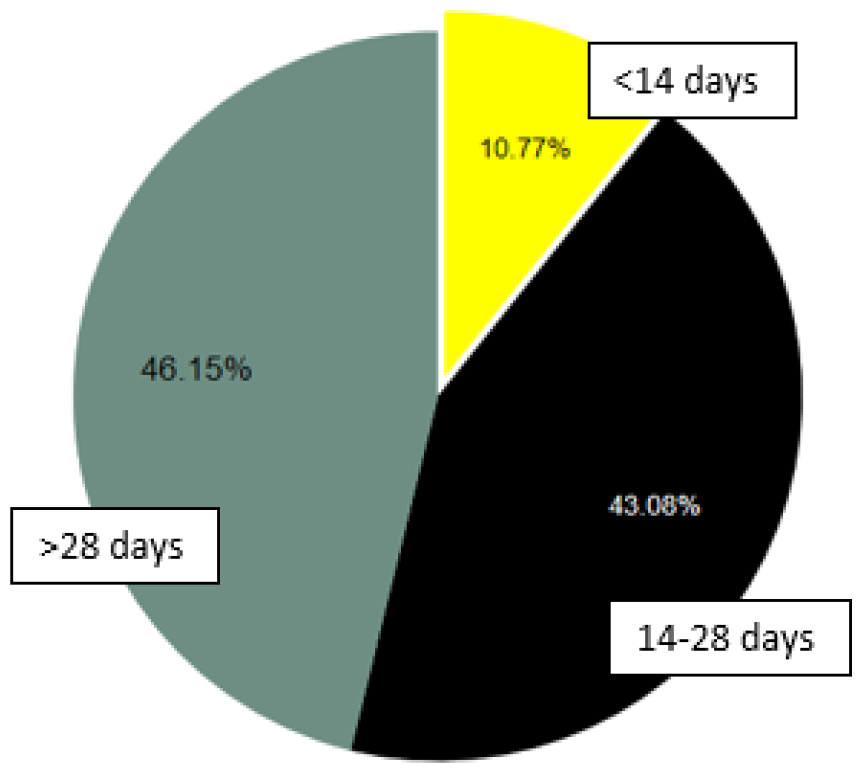
Pie chart illustrating lengths of hospitalization for TBM patients.

## Conclusion

This study showed a relatively high case fatality (49%) and mortality (2.22 per 100,000 per year) for Melanesian adults with Tuberculous Meningitis admitted to the Kundiawa General Hospital of Papua New Guinea. The case fatality for HIV-positive individuals with TB meningitis was significantly high at 93%.

90% of the patients stayed for more than 2 weeks and 46% stayed more than a month in the hospital.

28-day mortality in adult TB meningitis was strongly associated with HIV-TBM co-infection (p-value=0.007, 95% CI 2.49-289.19, and weakly associated with a positive fluid balance 24-hours after admission and admission GCS ≤ 10. Therefore, these parameters can serve as useful clinical predictors of mortality in adult TB meningitis patients in resource-constrained settings.

Prospective cohort studies of larger sample size are needed to further explore and ascertain the associations observed in this study.

## Data Availability

All data produced in the present study are available upon reasonable request to the authors

## Ethics

Ethical approval was granted by the interim Institutional Review Committee of the Simbu Provincial Health Authority-Clinical Research Center.

## Competing Interest Statement

The authors have declared no competing interest.

## Funding Statement

This study did not receive any funding

## Acknowledgement

I would like to sincerely thank the following individuals and institutions without whose help and support this study would not have been possible:

- My spouse and colleague Dr. Jacqueline Pahun
- Sir Joseph Nombri Memorial-Kundiawa General Hospital administration and ward staff: Dr. Francis Wandi, Dr. Raymond Saulep, Dr. Cassius Maingu, Dr. Kassi Ken, Dr. Izzard Agua, Resident Doctors and HEOs, Sr. Moro Drua and the Nursing Staff.
- Internal Medicine Department, UPNG SMHS & PGMH: Dr. Alexander Maha, Prof. David Linge, Prof. Sir Isi. H. Kevau, Dr. Will Toua.

## Abbreviations

TBM: Tuberculous Meningitis
TB: Tuberculosis
WHO: World Health Organization
GCS: Glasgow Coma Scale
CSF: Cerebrospinal Fluid
PCR: Polymerase Chain Reaction
CI: Confidence Interval
AFB: Acid Fast Bacilli
NGT: Nasogastric Tube
SiPHA: Simbu Provincial Health Authority
RCT: Randomized Control Trial.

## Notes

### Author Declarations

Ethical consideration and approval was granted by the Institutional Review Committee of the SiPHA-Clinical Research Center.

### Summary of Updates

Minor errors in graphs and language that have been updated to appropriation.

## References

1. World Health Organization, 2014. World Health Organization Global Tuberculosis Report 2013.

2. Thao, L.T.P., Heemskerk, A.D., Geskus, R.B., Mai, N.T.H., Ha, D.T.M., Chau, T.T.H., Phu, N.H., Chau, N.V.V., Caws, M., Lan, N.H. and Thu, D.D.A., 2018. Prognostic models for 9-month mortality in tuberculous meningitis. Clinical Infectious Diseases, 66(4), pp.523–532.

3. Chiang, S.S., Khan, F.A., Milstein, M.B., Tolman, A.W., Benedetti, A., Starke, J.R. and Becerra, M.C., 2014. Treatment outcomes of childhood tuberculous meningitis: a systematic review and meta-analysis. The Lancet Infectious Diseases, 14(10), pp.947–957.

4. Thwaites, G.E. and Hien, T.T., 2005. Tuberculous meningitis: many questions, too few answers. The Lancet Neurology, 4(3), pp.160–170.

5. Stadelman, A.M., Ellis, J., Samuels, T.H., Mutengesa, E., Dobbin, J., Ssebambulidde, K., Rutakingirwa, M.K., Tugume, L., Boulware, D.R., Grint, D. and Cresswell, F.V., 2020, August. Treatment outcomes in adult tuberculous meningitis: a systematic review and meta-analysis. In Open forum infectious diseases (Vol. 7, No. 8, p. ofaa257). US: Oxford University Press.

6. World Health Organization, 2007. Improving the diagnosis and treatment of smear-negative pulmonary and extrapulmonary tuberculosis among adults and adolescents: recommendations for HIV-prevalent and resource-constrained settings (No. WHO/HTM/TB/2007.379). World Health Organization.

7. Getahun, H., Harrington, M., O’Brien, R. and Nunn, P., 2007. Diagnosis of smear-negative pulmonary tuberculosis in people with HIV infection or AIDS in resource-constrained settings: informing urgent policy changes. The Lancet, 369(9578), pp.2042–2049.

8. Wen, L., Li, M., Xu, T., Yu, X., Wang, L. and Li, K., 2019. Clinical features, outcomes and prognostic factors of tuberculous meningitis in adults worldwide: systematic review and meta-analysis. Journal of neurology, 266(12), pp.3009–3021.

9. Li, K., Tang, H., Yang, Y., Li, Q., Zhou, Y., Ren, M., Long, X., Shen, W., Hu, R., Wang, X. and Zeng, K., 2017. Clinical features, long-term clinical outcomes, and prognostic factors of tuberculous meningitis in West China: a multivariate analysis of 154 adults. Expert review of anti-infective therapy, 15(6), pp.629–635.

10. Sunbul, M., Atilla, A., Esen, S., Eroglu, C. and Leblebicioglu, H., 2005. Thwaites’ diagnostic scoring and the prediction of tuberculous meningitis. Medical Principles and Practice, 14(3), pp.151–154.

11. Thwaites, G.E., Bang, N.D., Dung, N.H., Quy, H.T., Oanh, D.T.T., Thoa, N.T.C., Hien, N.Q., Thuc, N.T., Hai, N.N., Lan, N.T.N. and Lan, N.N., 2004. Dexamethasone for the treatment of tuberculous meningitis in adolescents and adults. New England Journal of Medicine, 351(17), pp.1741–1751.

12. Allen, B., 2014. Papua New Guinea national census 2011: Rates of population change in local level government areas.

13. Soria, J., Metcalf, T., Mori, N., Newby, R.E., Montano, S.M., Huaroto, L., Ticona, E. and Zunt, J.R., 2019. Mortality in hospitalized patients with tuberculous meningitis. BMC infectious diseases, 19(1), pp.1–7.

14. Hosoglu, S., Geyik M, F., Balik, I., Aygen, B., Erol, S., Aygencel T G., Mert, A., Saltoglu, N., Dokmetas, I., Felek, S. and Sunbul, M., 2002. Predictors of outcome in patients with tuberculous meningitis. The International Journal of Tuberculosis and Lung Disease, 6(1), pp.64–70.

15. Woldeamanuel, Y.W. and Girma, B., 2014. A 43-year systematic review and meta-analysis: case-fatality and risk of death among adults with tuberculous meningitis in Africa. Journal of neurology, 261(5), pp.851–865.

16. Smith, I., 2003. Mycobacterium tuberculosis pathogenesis and molecular determinants of virulence. Clinical microbiology reviews, 16(3), pp.463–496.

17. Davis, A.G., Rohlwink, U.K., Proust, A., Figaji, A.A. and Wilkinson, R.J., 2019. The pathogenesis of tuberculous meningitis. Journal of leukocyte biology, 105(2), pp.267–280.

18. Thee, S., Basu Roy, R., Blázquez-Gamero, D., Falcón-Neyra, L., Neth, O., Noguera-Julian, A., Lillo, C., Galli, L., Venturini, E., Buonsenso, D. and Götzinger, F., 2021. Treatment and Outcome in Children with Tuberculous Meningitis: A Multicenter Pediatric Tuberculosis Network European Trials Group Study. Clinical Infectious Diseases.

19. Garg, R.K. and Sinha, M.K., 2011. Tuberculous meningitis in patients infected with human immunodeficiency virus. Journal of neurology, 258(1), pp.3–13.

20. Thinyane, K.H., Motsemme, K.M. and Cooper, V.J.L., 2015. Clinical presentation, aetiology, and outcomes of meningitis in a setting of high HIV and TB prevalence. Journal of tropical medicine, 2015.

21. Cecchini, D., Ambrosioni, J., Brezzo, C., Corti, M., Rybko, A., Perez, M., Poggi, S. and Ambroggi, M., 2009. Tuberculous meningitis in HIV-infected and non-infected patients: comparison of cerebrospinal fluid findings. The International Journal of Tuberculosis and Lung Disease, 13(2), pp.269–271.

22. Brancusi, F., Farrar, J. and Heemskerk, D., 2012. Tuberculous meningitis in adults: a review of a decade of developments focusing on prognostic factors for outcome. Future microbiology, 7(9), pp.1101–1116.

23. Donovan, J., Rohlwink, U.K., Tucker, E.W., Hiep, N.T.T., Thwaites, G.E., Figaji, A.A. and Tuberculous Meningitis International Research Consortium, 2019. Checklists to guide the supportive and critical care of tuberculous meningitis. Wellcome Open Research, 4.

24. Harrigan, M.R., 2001. Cerebral salt wasting syndrome. Critical care clinics, 17(1), pp.125–138.

25. Ganong, C.A. and Kappy, M.S., 1993. Cerebral salt wasting in children: the need for recognition and treatment. American Journal of Diseases of Children, 147(2), pp.167–169.

26. Palmer, B.F., 2003. Hyponatremia in patients with central nervous system disease: SIADH versus CSW. Trends in Endocrinology & Metabolism, 14(4), pp.182–187.

27. Cotton, M.F., Donald, P.R., Schoeman, J.F., Van Zyl, L.E., Aalbers, C. and Lombard, C.J., 1993. Raised intracranial pressure, the syndrome of inappropriate antidiuretic hormone secretion, and arginine vasopressin in tuberculous meningitis. Child’s Nervous System, 9(1), pp.10–15.

28. Yasar, K.K., Pehlivanoglu, F. and Sengoz, G., 2010. Predictors of mortality in tuberculous meningitis: a multivariate analysis of 160 cases. The International journal of tuberculosis and lung disease, 14(10), pp.1330–1335.

29. Nataprawira, H.M., Ruslianti, V., Solek, P., Hawani, D., Milanti, M., Anggraeni, R., Memed, F.S. and Kartika, A., 2016. Outcome of tuberculous meningitis in children: the first comprehensive retrospective cohort study in Indonesia. The International Journal of Tuberculosis and Lung Disease, 20(7), pp.909–914.

30. Sheu, J.J., Yuan, R.Y. and Yang, C.C., 2009. Predictors for outcome and treatment delay in patients with tuberculous meningitis. The American journal of the medical sciences, 338(2), pp.134–139.

31. Khonga, M. and Nicol, M.P., 2018. Xpert MTB/RIF Ultra: a gamechanger for tuberculous meningitis? The Lancet Infectious Diseases, 18(1), pp.6–8.

